# General Health Checks in Adult Primary Care: A Review

**DOI:** 10.1101/2021.02.12.21251649

**Authors:** David T. Liss, Toshiko Uchida, Cheryl L. Wilkes, Ankitha Radakrishnan, Jeffrey A. Linder

## Abstract

**Importance:** General health checks—also known as general medical exams, periodic health evaluations, checkups, or wellness visits—to identify and prevent disease are extremely common in adult primary care. Although general health checks are often expected and advocated by patients, clinicians, payers, and health systems, others question their value. The current evidence was updated and recommendations provided for conducting general health checks in adults.

**Observations:** Randomized trials and observational studies with control groups from prior systematic reviews and an updated literature review through December 2020 were included. Out of 19 included randomized trials (906 to 59,616 participants; follow-up, 1 to 30 years), 5 evaluated a single general health check and 7 evaluated annual health checks. All of 12 included observational studies (240 to 471,415 participants; follow-up, cross-sectional to 5 years) evaluated a single general health check. General health checks were generally not associated with decreased mortality, cardiovascular events, or cardiovascular disease incidence. For example, in the South-East London Screening Study (n=7,229), adults age 40 to 64 who were invited to two health checks over two years experienced no 8-year mortality benefit (6% overall). However, general health checks were associated with increased detection of chronic diseases, such as depression and hypertension; moderate improvements in controlling risk factors such as blood pressure and cholesterol; increased clinical preventive service uptake, such as colorectal and cervical cancer screening; and improvements in patient-reported outcomes, such as quality of life and self-rated health. General health checks were sometimes associated with modest improvements in health behaviors such as physical activity and diet. For example, in the OXCHECK trial (n=4121), fewer intervention participants exercised less than once per month (68% versus 71%). Potential adverse effects in individual studies included an increased risk of stroke and increased mortality attributed to increased completion of advanced directives.

**Conclusions and Relevance:** General health checks were not associated with reduced mortality or cardiovascular events, but were associated with increased chronic disease recognition and treatment; risk factor control, preventive service uptake, and patient-reported outcomes. Primary care teams may reasonably offer general health checks, especially for groups at high risk of overdue preventive services, uncontrolled risk factors, low self-rated health, or poor connection to primary care.

## INTRODUCTION

General health checks – also known as general medical exams, periodic health evaluations, checkups, or wellness visits – are the second most common reason for ambulatory care visits in the United States (U.S.), accounting for about eight percent of the roughly 900 million annual U.S. ambulatory visits.^1^ Nearly all insurers provide partial or full financial coverage for these visits through mechanisms such as preventive visits for commercially insured patients^2^ and annual wellness visits for Medicare beneficiaries.^3^

Despite the prevalence of general health checks in everyday clinical care, questions persist about the value, goals, and components of these visits. For example, the Society of General Internal Medicine explicitly recommends *against* annual general health checks for asymptomatic adults.^4^ Others have called for these exams to be optimized so that physician-led teams deliver an “annual health review” that promotes trusting therapeutic relationships.^5^ An additional source of confusion is the term “annual physical,” a misnomer in that general health checks do not necessarily need to delivered annually nor include a physical examination beyond blood pressure and body mass index.^6^

### Prior Systematic Reviews

Prior reviews reached varying conclusions about the value of general health checks (**Table 1**). In 2007, an Agency for Healthcare Research and Quality (AHRQ) Evidence Report/Technology Assessment^7^ and accompanying review manuscript^8^ found that periodic health evaluations were consistently associated with receipt of some cancer screenings and cholesterol screening, and could reduce patient worry. However, periodic health evaluations had mixed effects on mortality, other clinical outcomes, and costs. In 2016, a Cochrane review of systematic risk assessment for primary prevention of cardiovascular disease^9^ found no effects on mortality or cardiovascular endpoints, but some evidence of reductions in total cholesterol and blood pressure. In 2019, a separate Cochrane review of general health checks in adults^10^found little or no effect on total mortality, cancer mortality, or ischemic heart disease, and probably little or no effect on cardiovascular mortality or stroke. The three systematic reviews described the risk of bias in included studies as generally variable, high, or unclear.

**Table 1:** Characteristics of prior systematic reviews.

| Review Topic | Included Study Designs | Inclusion & Exclusion Criteria | Outcomes | Description of Included Trials | Locations of Included Trials (n) | Main Findings |
| --- | --- | --- | --- | --- | --- | --- |
| Periodic health evaluations<sup>7,8</sup> | Randomized trials and observational studies | Attended visit assessing overall health and risk factors for preventable disease. Excluded studies where all patients under age 18. | Clinical preventive services, clinical outcomes, and costs | 10 trials published between 1973 and 1999<br>Participant range, 112 to 32,186 | *Asia* (1): Japan (1)<br>*Europe* (2): Sweden (1), U.K. (1)<br>*North America* (7): Canada (1), U.S. (6) | PHEs associated with receipt of gynecologic exams, Papanicolaou smears, cholesterol screening, and FOBT (medium or high quality of evidence). PHEs reduced patient worry (medium quality of evidence), but had mixed effects on other clinical outcomes and costs. |
| Systematic risk assessment for primary prevention of CVD<sup>9</sup> | Randomized trials | Healthy adults, including those at moderate or high CVD risk. Excluded studies of patients with known CVD. | *Primary:* all-cause and cardiovascular mortality, non-fatal cardiovascular endpoints<br>*Secondary:* major CVD risk factors, intermediate program outcomes, harms, costs | 9 trials published between 1980 and 2014<br>Participant range, 501 to 145,441 | *Europe* (6): Denmark (2), Sweden (1), U.K. (2), multiple countries (1)<br>*North America* (3): Canada (1), U.S. (2) | Systematic risk assessment had no effect on mortality or cardiovascular endpoints (low quality of evidence). Some favorable effects on total cholesterol, systolic blood pressure, and diastolic blood pressure (low or very low quality of evidence). |
| General health checks in adults<sup>10</sup> | Randomized trials | Adults in a primary care or community setting. Excluded studies targeting older adults and populations with | *Primary:* total mortality and disease-specific mortality | 15 trials published between 1965 and 2017 | *Europe* (12): Denmark (3), Sweden (4), U.K. (4), multiple | GHCs had little or no effect on total mortality, cancer mortality, or ischemic heart disease (high certainty of evidence). Probably |
|  |  | specific risk factors/ conditions. | *Secondary:* several, including cardiovascular events | Participant range, 533 to 61,301 | countries (1)<br>*North America* (3): U.S. (3) | little or no effect on cardiovascular mortality or stroke (moderate certainty of evidence). |

Abbreviations: FOBT, fecal occult blood testing; PHE, periodic health evaluation; U.K., United Kingdom; U.S., United States

However, prior review findings may have limited generalizability to modern primary care populations. Prior reviews differed with regard to exclusion^10^ or inclusion^8^ of trials in elderly patients (**Table 1**), thus omitting U.S.-based randomized trials among Medicare enrollees.^11-14^ Many trials were conducted in European countries with “usual care” consisting of universally insured populations with population-level access to primary care and few cost-related access barriers.^15^ Additionally, many trials were conducted before several modern pharmacotherapies—including statins, many antihypertensive medications, and smoking cessation medications—were available. Findings from prior reviews may not be generalizable to U.S. primary care sthe ettings.^16^

The literature on general health checks was reviewed to gain an up-to-date understanding of the evidence. In addition to considering results of randomized trials, controlled, observational evidence from the twenty-first century was also evaluated.

## METHODS

Complementary approaches were used to identify randomized trials (regardless of publication year) and observational studies (published between January 2000 and November 2019) that met inclusion criteria. In addition to considering all randomized trials included in prior systematic reviews;^8-10^ we also considered observational studies from the prior review by Boulware et al.^8^ that were published in 2000 or later. In addition, the Ovid MEDLINE database was queried on January 19, 2021 to identify articles published up to December 31, 2020. The MEDLINE query included three related searches (Supplement). First, a search for observational articles published between 2000 and 2020 was conducted. Then, to identify recently published randomized trials, the search strategy from each of the two prior Cochrane reviews^9,10^ was replicated, but restricted to articles published since each review’s respective search date. One study author screened each study abstract; abstracts identified as potentially meeting inclusion criteria were then screened by an additional author. If both screeners agreed that the study might meet inclusion criteria, two authors then screened the full text manuscript. When an author pair disagreed about whether to include a study in the review, the full study team screened the relevant manuscript and jointly made a final inclusion decision. Reference lists of included studies were also hand searched for potentially eligible articles.

The review included controlled, randomized trials and controlled (i.e., with a comparator group), observational studies conducted in primary care among adults—with no upper age limit—with a total sample size of at least 200 patients to eliminate small studies that would be more likely to provide imprecise results. Disease-specific or condition-specific studies and studies conducted in settings outside of primary care practices, such as patients’ homes, workplaces, or freestanding pharmacies were excluded. Articles that were not available in English and studies where outcome data were not yet published were excluded.

To be included, studies had to report outcome measures from at least one of the following seven categories: 1) mortality; 2) cardiovascular outcomes, including cardiac events, such as stroke, or incidence of conditions such as coronary heart disease or diabetes; 3) chronic disease detection, defined by a newly diagnosed chronic illness or initiation of chronic disease treatment (typically via pharmacotherapy); 4) risk factor control for outcomes such as weight, blood pressure, cholesterol, or overall cardiovascular risk; 5) uptake of clinical preventive services; 6) health behaviors such as exercise, diet, or smoking, and; 7) patient-reported outcomes such as quality of life or anxiety. For each included study, relevant data on these seven outcome categories were collected, along with any available data on potential adverse effects among patients exposed to general health checks.

## OBSERVATIONS

After screening 1,860 abstracts, a total of 31 studies met inclusion criteria: 19 were randomized trials and 12 were controlled observational studies (**Figure 1**).

**Figure 1:**
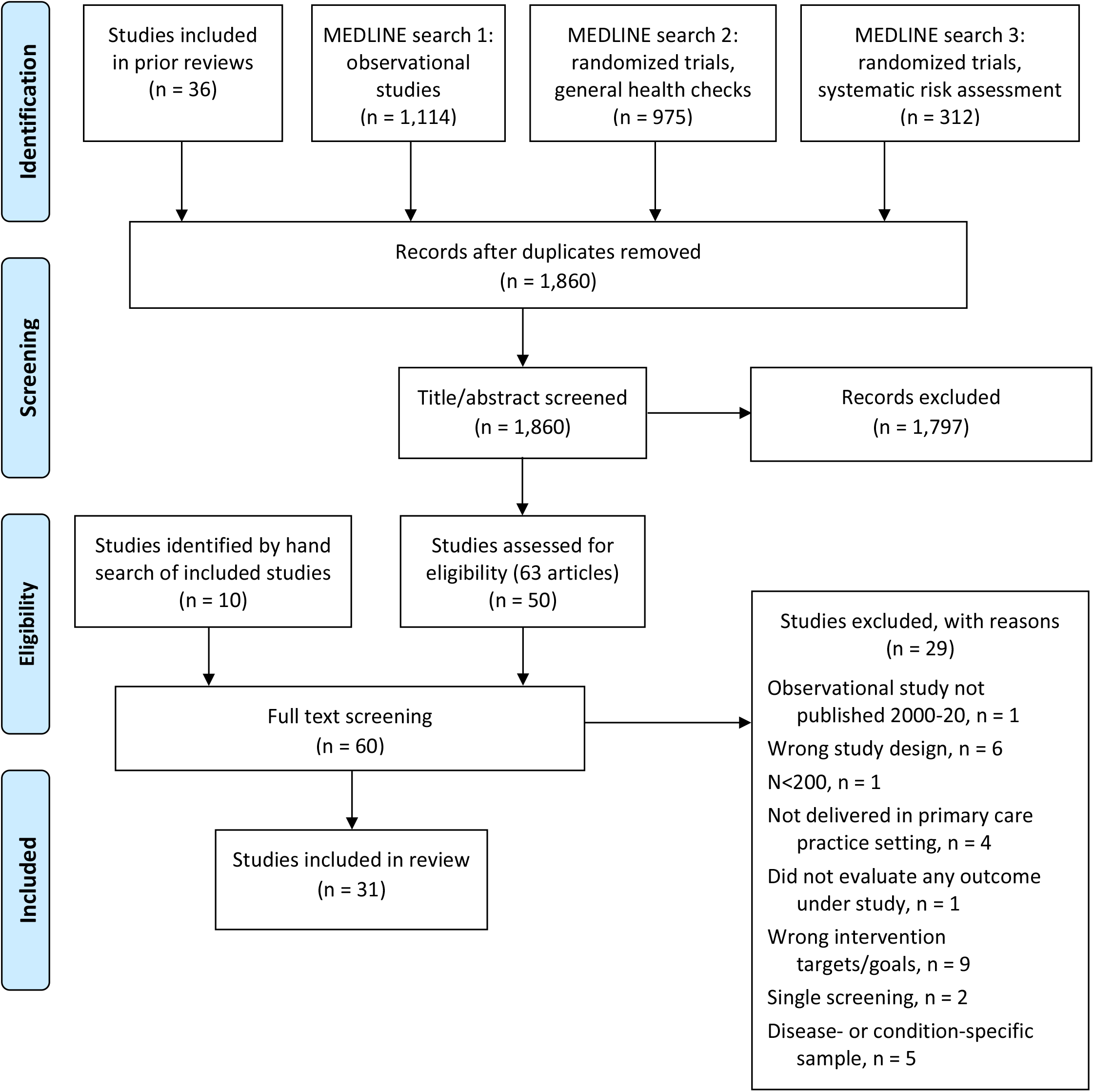
Summary of Evidence Search and Selection.

The studies were conducted in multiple countries, in different age groups, and varied in size. Seventeen studies (55%) were conducted in Europe (9 in Scandinavia, 8 in the United Kingdom [U.K.]), 11 (36%) in the U.S., and the remainder in Asia (6%) or Canada (3%). Six studies (19%) were restricted to patients between age 40 and 64, eight (26%) were restricted to patients age 65 and older, and 17 (55%) included patients in multiple or unspecified age groups. The number of included study subjects in randomized trials ranged from 906 to 59,616 and, in observational studies, from 240 to 471,415. Follow-up periods ranged from 6 months to 30 years. Fourteen included studies (45%) began prior to 1990 (i.e. before medications such as statins were widely available), 7 (23%) began during the 1990s, and 10 (32%) began during the twenty-first century.

Although a physician delivered at least some components of all included interventions, general health check intervention formats and components varied widely. The primary care physician conducted a physical examination in 16 (52%) included studies. Some general health check interventions included a single physician-led visit, while others included a multiphasic screening session with a non-physician, followed by a visit with a physician to review screening results. Many different forms of screening were employed, including interviews to assess patient-reported risk factors, laboratory testing of blood and urine, cancer screenings, and other diagnostic testing of lung, heart, or eye function. Lifestyle interventions and health behavior coaching sometimes accompanied general health check screenings.

The frequency of general health checks varied by study design. While all 12 included observational studies evaluated a single general health check, there was substantial variation across included randomized trials. Five of 19 (26%) trials evaluated a single, comprehensive general health check, 7 (37%) evaluated annual health checks, 1 (5%) evaluated biaannual health checks, and 6 (32%) evaluated health checks delivered at other frequencies.

### Mortality

General health checks were not consistently associated with changes in mortality. In 13 randomized trials that evaluated mortality (**Table 2**),^12,17-28^ eleven reported no significant all-cause mortality benefit.^11,12,17-19,21-23,25-27^ For example, in the South-East London Screening Study (n=7,229), adults age 40 to 64 who were invited to two multiphasic screening sessions over two years experienced no benefit in eight-year mortality (6% overall)^21^ or nine-year mortality (results not reported).^29^ In the DanMONICA trial conducted among 17,845 Danes, invitation to screenings and interventions at baseline, five years, and 10 years led to no difference in 30-year all-cause mortality.^25^

**Table 2:**
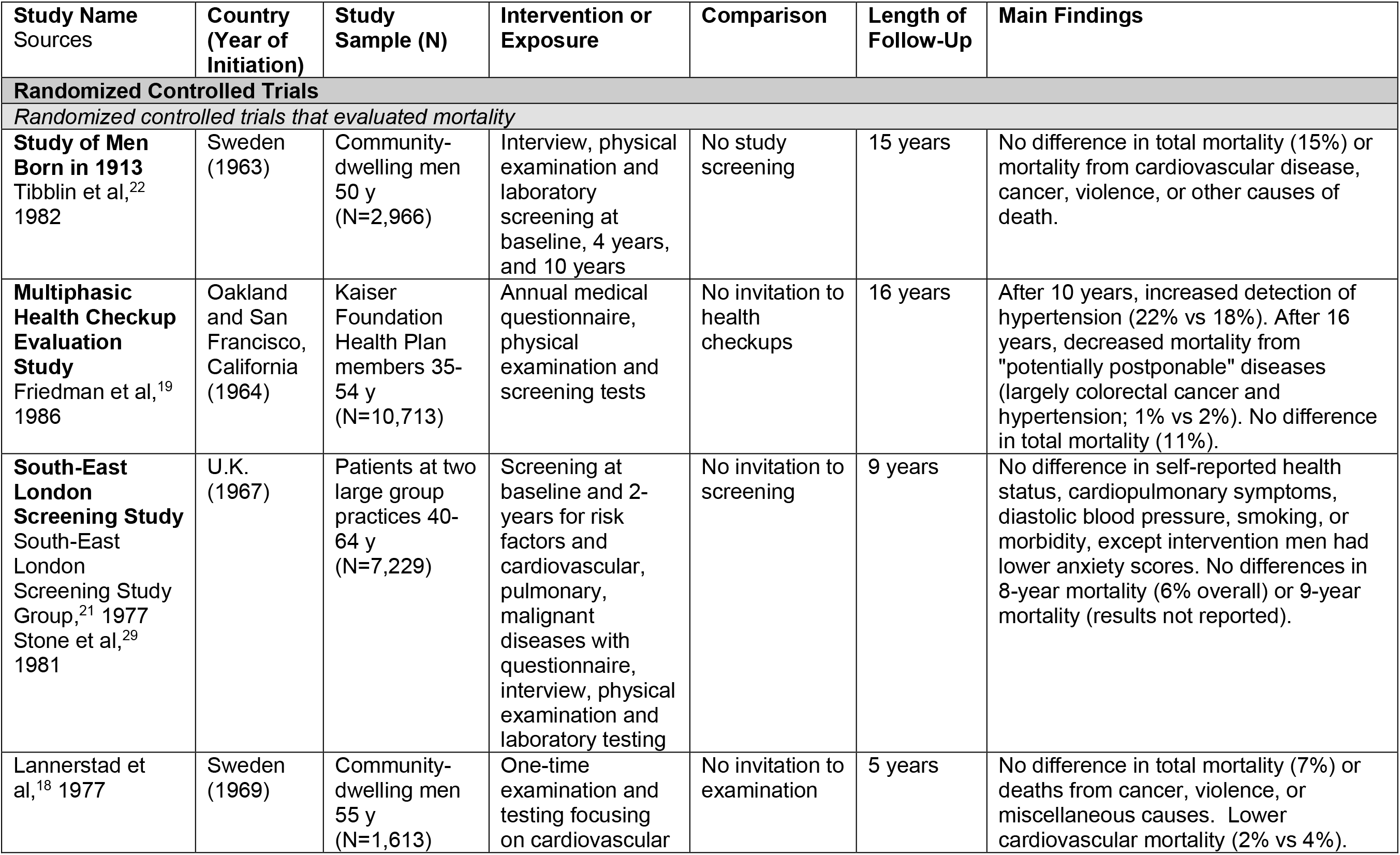
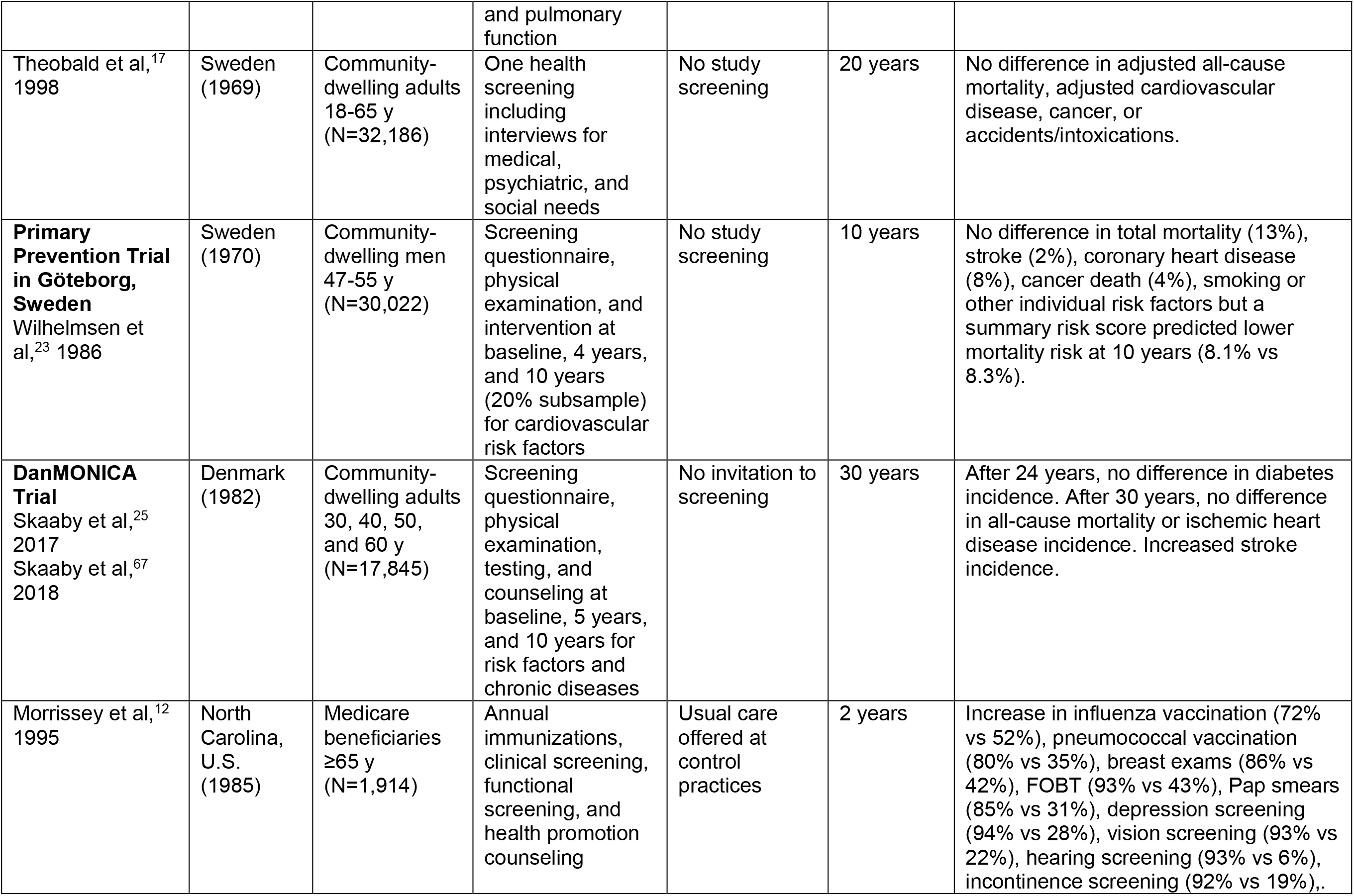
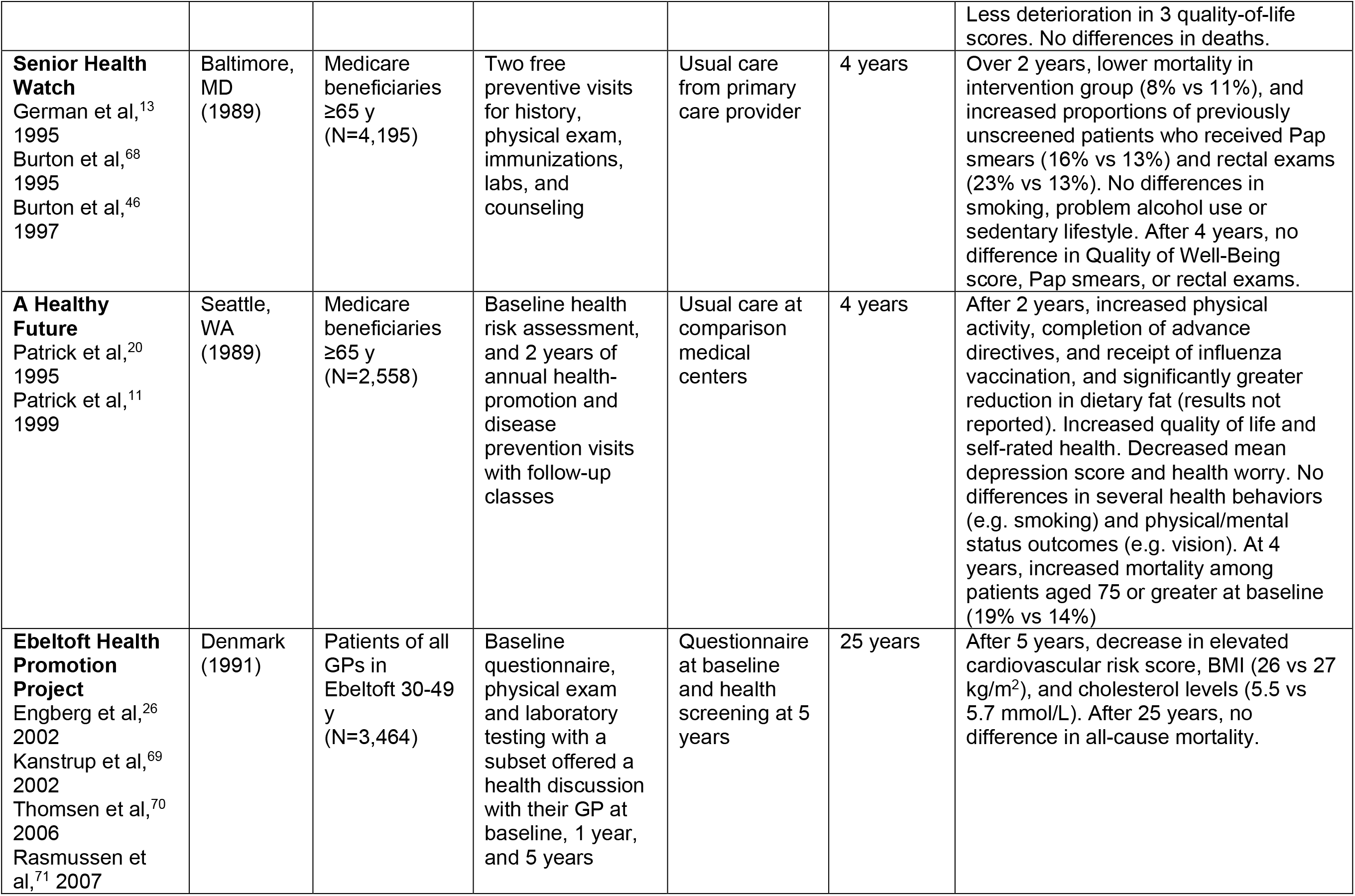
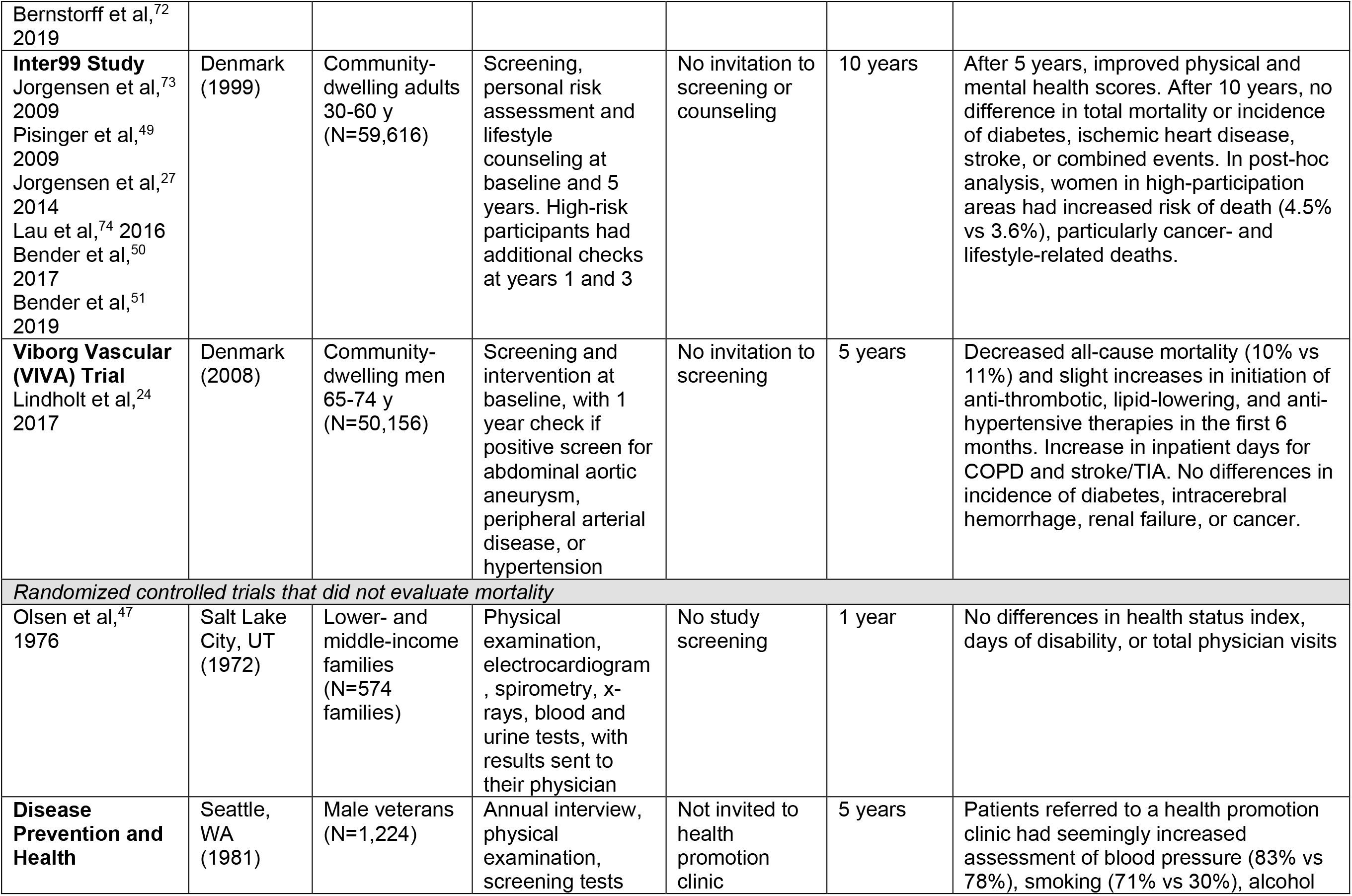
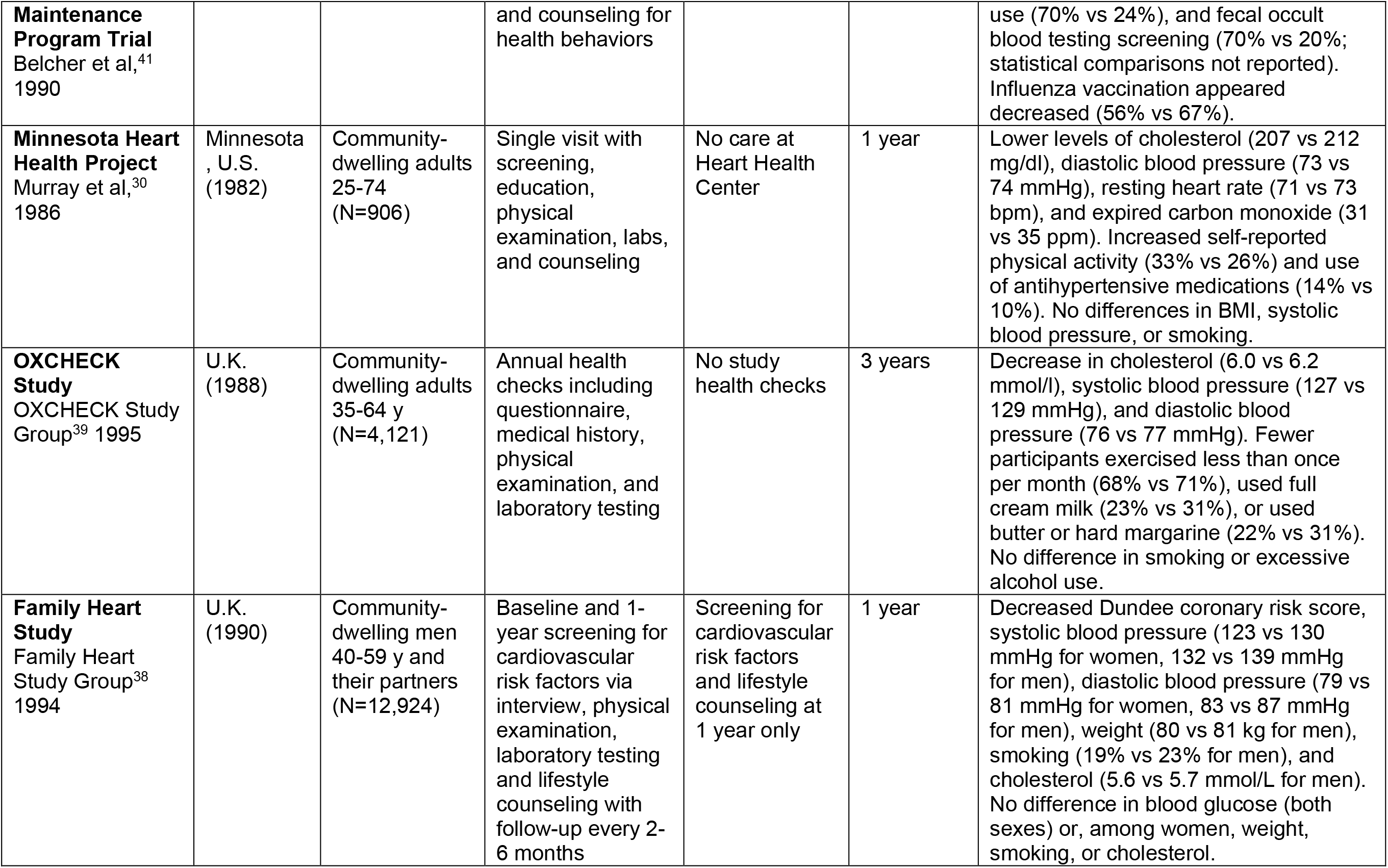
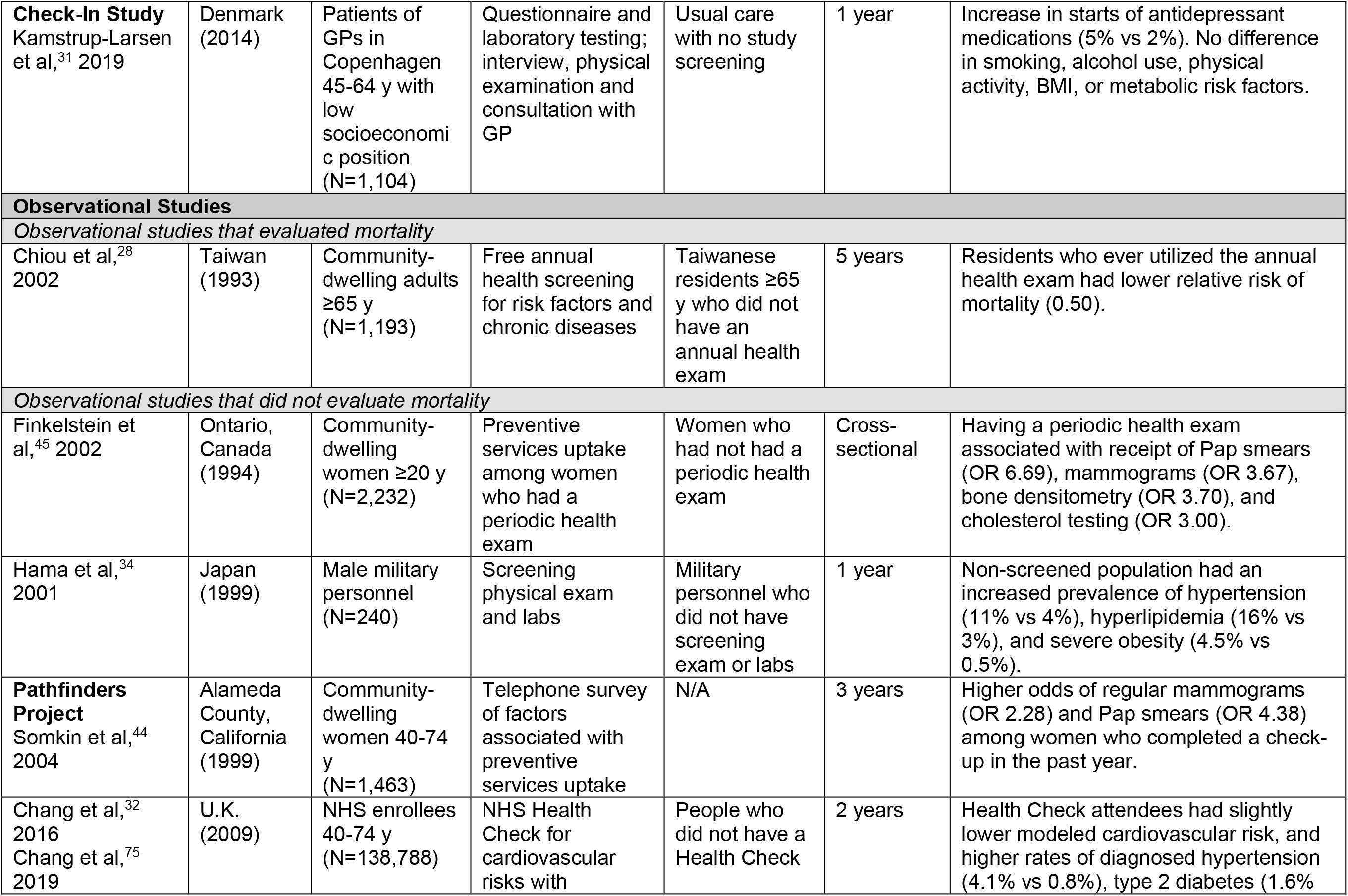
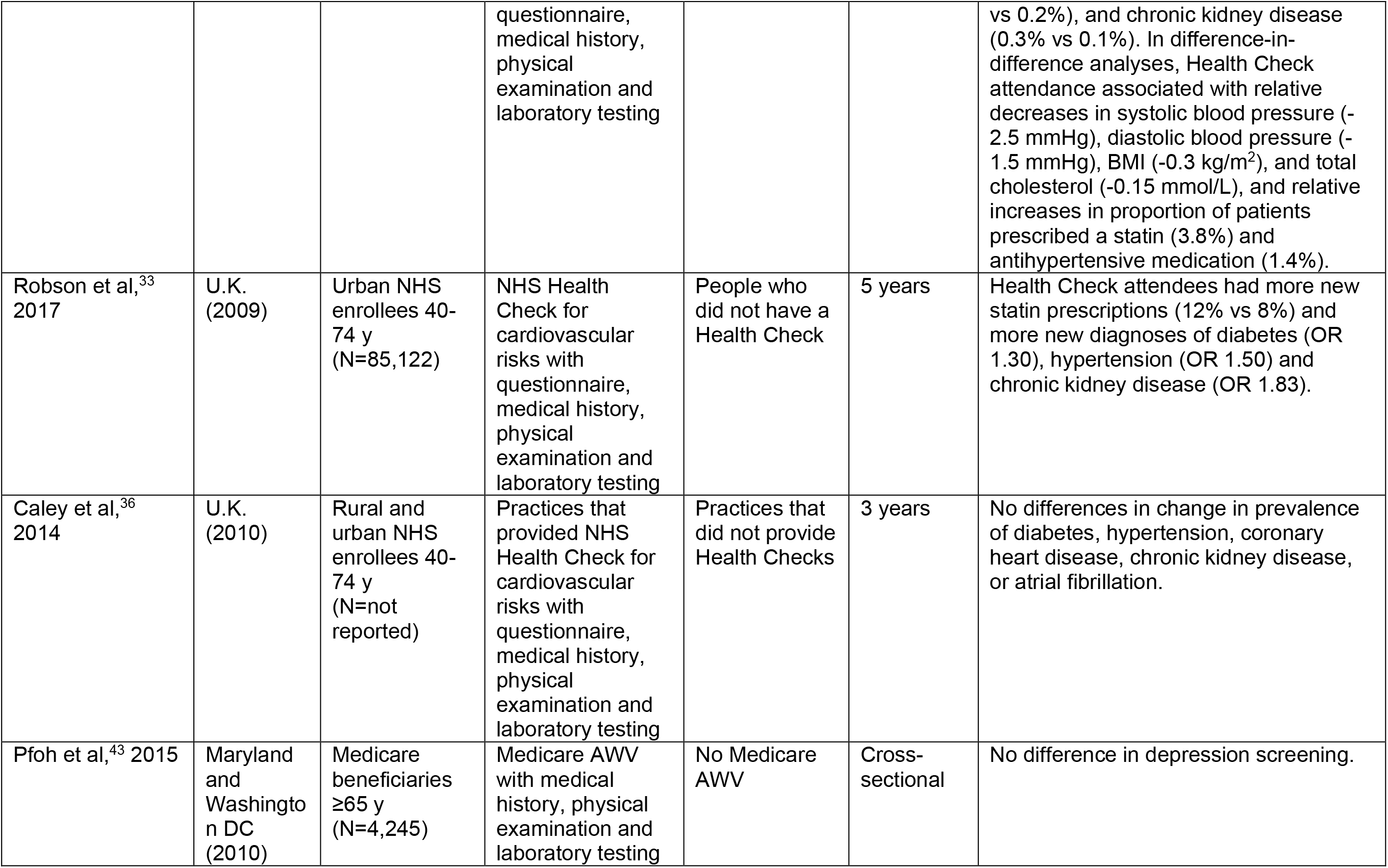
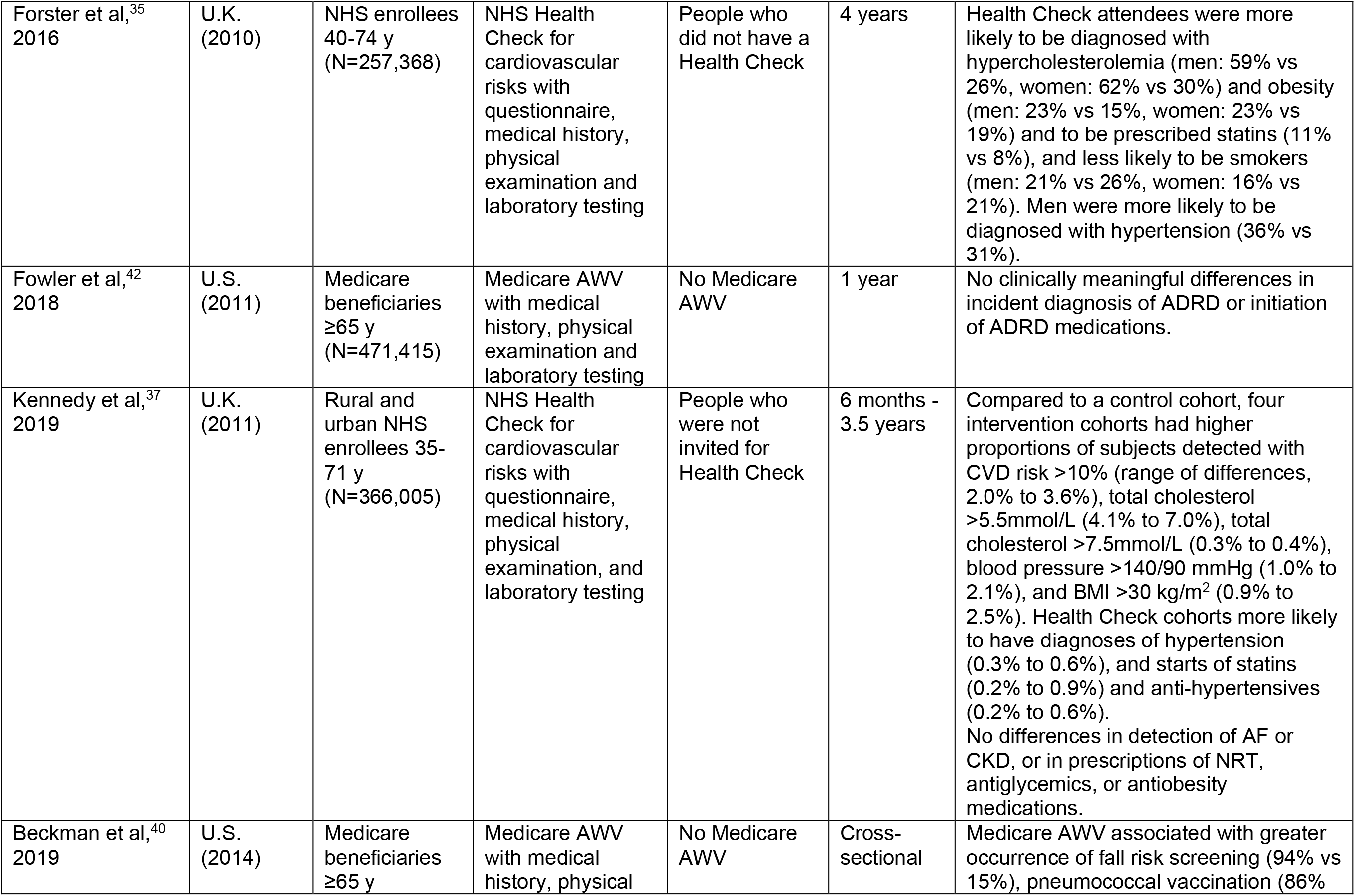
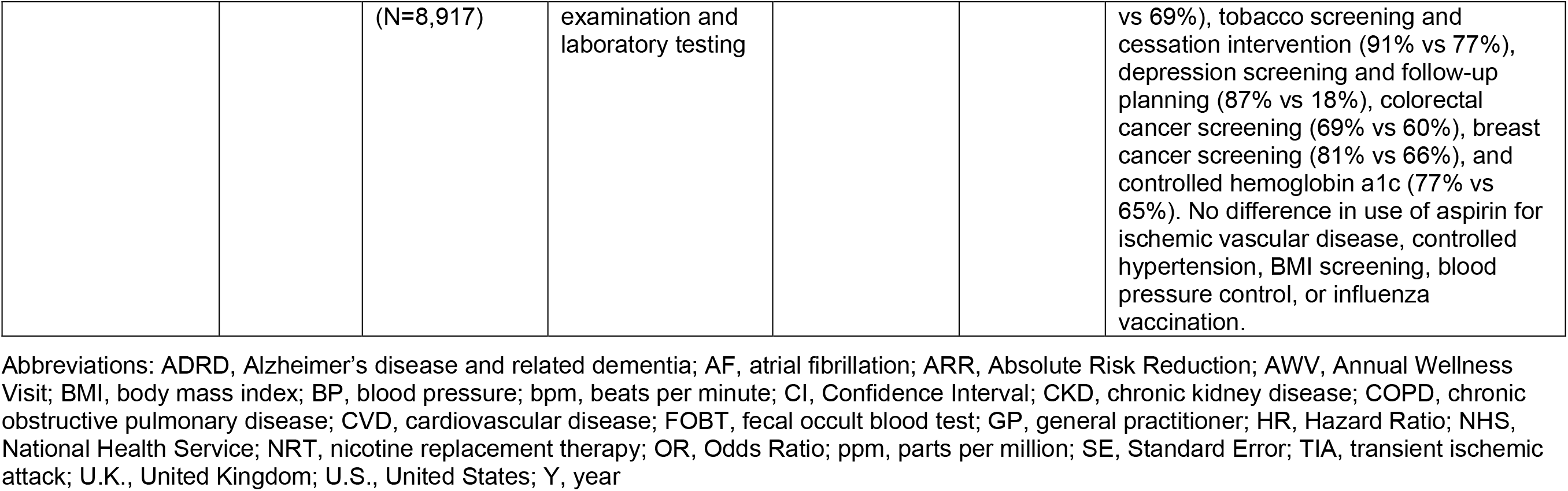
Summary of included studies, by study design and whether mortality evaluated.

General health checks showed a mortality benefit in two trials conducted in older adults. In the Senior Health Watch trial (n=4,195), Medicare beneficiaries were offered two annual preventive visits including a comprehensive exam, laboratory procedures, and immunizations, plus physician-recommended follow-up counseling visits.^13^ Mortality was lower in the intervention group during the two-year intervention period (8% versus 11%^13^) and the four-year period including two years of post-intervention follow-up (19% versus 22%; P=0.02 in chi-square test conducted by our study team in Stata, version 14.2 [StataCorp; College Station, TX]). In the Viborg Vascular (VIVA) trial (n=50,156), Danish men aged 65 to 74 received a universal screening intervention—including portable ultrasound screening for abdominal aortic aneurysm, portable doppler screening for peripheral arterial disease, and hypertension screening. Over a median of 4.4 years follow-up, 10% of patients in the screening group died, compared to 11% of the non-screening group.^24^

### Cardiovascular Outcomes

In five included studies^23-27^—all randomized trials—that evaluated cardiovascular outcomes, general health checks consistently failed to reduce cardiovascular events or cardiovascular disease incidence. In the Inter99 trial (n=59,616), where Danish adults aged 30 to 60 received up to four health checks over five years, there were no differences in 10-year incidence of ischemic heart disease or stroke.^27^ Similarly, in the DanMONICA trial, there were no differences in 30-year ischemic heart disease incidence.^25^

### Chronic Disease Detection

In four randomized trials^19,24,30,31^ and six observational studies,^32-37^ detection of chronic disease was increased among patients receiving general health checks. In the recent Check-In Study trial among patients aged 45 to 64 with low levels of education, 5% of patients randomized to a single preventive health check, and 2% of those randomized to usual care, received a new antidepressant prescription over one year. However, there were no differences in detection of hypertension, hypercholesterolemia, or diabetes.^31^ In a matched case-control study of patients from ethnically diverse areas of London with high levels of social deprivation (n=85,122), attending a National Health Service (NHS) Health Check was associated with higher odds of newly diagnosed diabetes (odds ratio [OR] 1.30), hypertension (OR 1.50), and stage 3-5 chronic kidney disease (OR 1.83). Additionally, 12% of NHS Health Check attendees received a new statin prescription, versus 8% in patients who did not attend a health check.^33^

### Risk Factor Control

In 11 studies,^21,23,26,30-32,34,37-40^ 7 of which were trials, general health checks were associated with small or moderate improvements in measures such as blood pressure, cholesterol, and cardiovascular risk scores. In the Family Heart Study conducted in the U.K. (n=12,924), both women and men randomized to a general health check followed by tailored follow-up had reduced systolic blood pressure (6.4 mmHg reduction for women; 7.4 mmHg for men) and diastolic blood pressure (2.7 mmHg for women; 3.3 mmHg for men) after one year.^38^ Similarly, in the Minnesota Heart Health Project (n=906), where rural adults aged 25 to 74 were randomized to a single general health check visit including multimodal screening and education, the intervention group had a limited, but statistically significant, reduction in mean diastolic blood pressure (1.3 mmHg) and total cholesterol (4.6 mg/dl) after one year.^30^

General health checks had limited effect on weight. In the Family Heart Trial, mean weight among men in the intervention group was reduced by 1.2 kilograms (kg) after one year, and there were no weight differences between groups among women.^38^ In the Ebeltoft Health Promotion Project trial (n=3,464), patients in a rural Danish area who were offered general health checks in two successive years had lower mean body mass index (BMI) at five-year follow-up than controls (26 vs 27 kg/m^2^).^26^ However, in both the Minnesota Heart Health Project and the Check-In Study trial, a single general health check had no effect on BMI at one year.^30,31^

### Clinical Preventive Services

In four randomized trials^12,13,20,41^ and five observational studies^40,42-45^—all of which were conducted in North America—patients receiving general health checks frequently had higher uptake of clinical preventive services. For example, in a propensity score-matched analysis of 2015 data from a U.S. Accountable Care Organization (n=8,917), a higher proportion of patients who received a Medicare Annual Wellness Visit (AWV) had completed colorectal cancer screening (69% vs 60%), breast cancer screening (81% vs 66%), fall risk screening (94% vs 15%), pneumococcal vaccination (86% vs 69%), tobacco screening and cessation intervention (91% vs 77%), and depression screening and follow-up planning (87% vs 18%). However, there were no differences in the proportion of patients using aspirin for ischemic vascular disease or completing influenza vaccination.^40^

Across included studies, there was variation in the persistence of preventive service increases after patients stopped attending general health check visits. In the A Healthy Future trial (n=2,558), where Medicare beneficiaries received a preventive services benefit package over two years, the intervention arm experienced relative increases in proportions of patients receiving influenza vaccination and completing advance directives at both two-year and four-year follow-up (data not reported).^11^ In contrast, the Senior Health Watch trial produced increases in the proportions of patients who were unscreened at baseline who received Pap smears (16% vs 13%) and rectal exams (23% vs 13%) during the two-year period when annual preventive visits were offered free of charge, but not in subsequent years.^46^

### Health Behaviors

Across nine randomized trials^13,20,21,23,26,30,31,38,39^ and two observational studies,^32,35^ some studies demonstrated an association between general health checks and modest improvements in health behaviors such as exercise-and diet-related outcomes. In the Oxford and Collaborators Health Check (OXCHECK) trial (n=4,121), where U.K. adults aged 35 to 64 were randomized to annual health checks over three years, fewer participants reported exercising less than once per month (68% vs 71%) and use of butter or hard margarine (22% vs 31%), but there were no differences in excessive alcohol consumption.^39^ In the A Healthy Future trial, the intervention arm had greater improvements in physical activity and larger reductions in dietary fat after two years; however, these differences did not persist over four-year follow-up.^11^

General health check interventions generally did not reduce smoking rates. Although the proportion of smokers was lower among men in the Family Heart Trial’s intervention arm (19% vs 23%) after one year, there was no significant difference among women.^38^ Many other trials demonstrated no reductions in smoking.^20,23,26,30-32,39^

### Patient-Reported Outcomes

Of six randomized trials^12,13,20,21,27,47^ evaluating patient-reported outcome measures, five produced one or more significant findings.^12,13,20,21,27^ In three U.S.-based trials in Medicare beneficiaries, after two-year intervention periods the intervention group reported relative improvements in outcomes such as health worry,^11^ quality of well-being,^12,13^ quality of life,^11,12^ and self-rated health.^12^ In two European trials, general health check interventions separately led to reduced anxiety (among men)^48^ and improved self-rated health.^49^ In the only included trial with null findings for patient-reported outcomes, a multiphasic screening exam and physician follow-up produced no change in patient-reported health status among low- and middle-income families in Salt Lake City, UT.^47^

### Potential Adverse Effects

Four trials demonstrated potential adverse effects of general health checks.^20,24,25,27^ In the A Healthy Future trial testing a preventive services benefit package for Medicare beneficiaries, intervention arm participants aged 75 or greater had increased mortality over four years (19% vs 14%).^11^ A follow-up analysis attributed this finding to the intervention group’s increased completion of advance directives and decreased receipt of unwanted life-sustaining treatment in the face of serious medical events.^20^

In the Inter99 study—a population-based ischemic heart disease prevention trial in Denmark^27^ (n=59,616)—women in the intervention group who lived in high-participation communities had 32% higher all-cause mortality risk than controls (hazard ratio [HR] 1.32),^50^ which was driven by higher risks of lifestyle-related (HR 1.37) and cancer-related (HR 1.47) mortality. Study investigators had no explanation for this mortality increase, but hypothesized in a post-hoc analysis^51^ the potential for high use of nutritional supplements that could have increased smoking-related cancers.^52^

In the DanMONICA trial, patients in the intervention group—who were invited to as many as three health checks between 1982 and 1994—had 14% higher stroke incidence over 30-year follow-up.^25^ The authors hypothesized that this increase in strokes could be due to overdiagnosis, overtreatment, injury from testing, distress from test results, or false reassurance.

In the VIVA trial—in which mortality declined in the intervention group—of screening for abdominal aortic aneurysm, peripheral arterial disease, and hypertension among Danish menaged 65 to 74, patients in the intervention group spent more days in the hospital for inpatient admissions related to COPD (incidence rate ratio [IRR] 1.13) and stroke or transient ischemic attack (IRR 1.05). However, treatments received during these inpatient stays may have contributed to the mortality reductions in the intervention group.^24^

## DISCUSSION

General health checks are not associated with reduced mortality or cardiovascular outcomes. General health checks are associated with increases in chronic disease detection, moderate improvements in risk factor control, increased uptake of clinical preventive services, and improvements in patient-reported outcomes. Despite being associated with limited changes in some health behaviors, general health checks were not associated with reductions in smoking. General health checks may have possible adverse effects. Benefits were frequently observed during the active intervention period or within a few years of general health check attendance; benefits were generally not observed more than five years after completion of an intervention.

Some findings from this review mirror those of prior reviews, such as the observed lack of mortality benefit,^7,9,10^ increased clinical preventive service uptake,^8^ and limited improvements in risk factor control.^9^ However, in contrast to prior reviews, general health checks were found to improve patient-reported outcomes besides patient worry, with five of six included trials demonstrating benefits in outcomes such as quality of life and self-rated health.^12,13,20,21,27^ These findings are particularly notable in light of the demonstrated association between self-rated health and mortality risk.^53,54^ Additionally, this review included seven randomized trials from the U.S., more than any prior review.^8-10^ The seven included U.S.-based trials generally produced positive findings, particularly for clinical preventive services and patient-reported outcomes. Given that the U.S. is unique among high-income countries in its lack of universal insurance and the access barriers faced by patients, it may be reasonable to conclude that general health checks—by providing some of the benefits of available, accessible primary care^55^—are particularly effective in U.S. settings.

The Medicare AWV is one of the most common general health checks in the U.S. Although prior studies have produced mixed evidence on the potential benefits of AWVs,^40,56^ the current review bolsters the case for several services covered within AWVs,^57^ such as collection of patient-reported health status, assessment and review of risk factors, and updating a written schedule of recommended preventive services. However, not all AWV-covered services are necessarily beneficial. Cognitive screening is not recommended by the U.S. Preventive Services Task Force (USPSTF),^58^ and an observational study included in this review detected no association between AWV uptake and incident diagnosis of Alzheimer’s disease and related dementia.^42^ There are many unanswered questions about AWVs, such as whether annual AWVs confer greater benefits than less frequent AWVs.

Although many patients consider “the annual physical” necessary,^59^ general health checks do not necessarily need to occur every year or include a physical exam. Although several studies in this review evaluated annual general health check delivery, these studies did not clearly demonstrate that health checks need to be offered this frequently. In addition, beyond blood pressure measurement, BMI assessment, and Pap smears for women, a regular screening physical examination has not been shown to improve health.^6^ Given general health checks’ potentially limited but heterogeneous benefits across multiple domains, it seems rational to target populations with many preventive care needs or those at highest risk. For example, patients age 50 to 59 may be likely to benefit from general health checks because of the many USPSTF-recommended preventive services in this age group.^58^ General health checks may also be especially beneficial for the increasingly large number of U.S. adults with no recent primary care physician visits,^60^ and those at risk of high blood pressure, high cholesterol, low rates of preventive service uptake, or low reported quality of life or self-rated health. General health checks may be particularly beneficial in historically underserved populations, and have the potential to decrease health care disparities.^31,33^

Current evidence provides no clear “blueprint” for how to systematically deliver general health checks in modern primary care settings. The frequency, number, and content of health checks varied substantially among studies (Table 2, ‘Intervention or Exposure’ column), so it remains unclear how many, or how often, general health checks should be delivered. However, intervention components should employ efficient—and likely team-based—approaches to address USPSTF A and B recommendations, recommended immunizations,^61^ and ongoing risk factor control in diagnosed chronic illnesses. When providing general health checks, care teams should explicitly avoid delivery of non-recommended, low-value services. Depending on organizational resources and patient population risks, general health checks could also include screening and tailored support for patients’ social needs,^62^ or allotted time for conversations to promote trusted, healing relationships.^5^ Preventive services might be delivered “opportunistically,” outside of dedicated general health check visits. The COVID-19 pandemic has forced a reconsideration of traditional in-person care processes. Research should test new forms of general health checks that pair virtual components (e.g. telemedicine and patient portals) with brief in-person visits for services that cannot be delivered remotely (e.g. vaccinations). Additionally, pandemic-induced reductions in outpatient utilization^63^ could exacerbate the underuse of recommended preventive services, while increasing rates of undiagnosed chronic illness. As such, there is now a need—and opportunity—for health systems to collaborate with patients to design and test new care delivery processes for general health checks that rely on minimal amounts of in-person services.

This review has several limitations. First, unlike some prior reviews,^9,10^ this review was not a meta-analysis, precluding our ability to calculate pooled effect sizes. Second, the “dose” of general health checks–both frequency and duration–was often quite low. Third, despite our findings that general health checks were associated with improvements in multiple outcome domains, the benefits were sometimes quite small, particularly for risk factors such as blood pressure and cholesterol and health behaviors such as physical activity and diet. Fourth, general health check uptake is often high among relatively advantaged groups^45,64^ that are likely to be healthy or already have conditions diagnosed in the absence of general health checks; this selection bias is likely present in included observational studies. However, there were some included studies where general health check attendance was associated with increased chronic disease detection and treatment among disadvantaged groups, such as those with low education and inner-city residents.^31,33^ Fifth, the pragmatic nature of the evidence under study sometimes hindered our ability to isolate effects of general health check uptake in routine clinical practice settings. For example, in the Multiphasic Health Checkup Evaluation Study, patients randomized to the intervention attended a mean of seven general health checks over 16-year follow-up, versus three general health checks in the usual care group, thus producing a comparison of varying general health check exposures (but not a truly unexposed control group). Also, in the DanMONICA trial, although up to three general health checks over 10 years did not reduce 30-year mortality or ischemic heart disease incidence, it is unclear whether these findings should be attributed to the ineffectiveness of general health checks, “opportunistic” delivery of recommended preventive services outside of dedicated general health check visits,^65^ or an intervention of insufficient duration. Sixth, general health checks’ potential adverse effects and costs have not been evaluated as fully as potential benefits. Seventh, included studies did not examine the relational benefits of a regular general health check.^5^ Finally, there is a dearth of recent high-quality evidence. The majority of included trials were conducted at least 20 years ago, and most large studies were conducted prior to the introduction of statins, which are known to reduce outcomes such as blood cholesterol and stroke risk.^66^

## CONCLUSION

General health checks have no effect on mortality or cardiovascular outcomes, but are associated with increased chronic disease detection and preventive service uptake, improvements in patient-reported outcomes, and limited improvements in risk factor control and some health behaviors. Primary care practices and health systems that elect to offer general health checks should target groups of patients that can substantially benefit, and should investigate new approaches to maximizing general health checks’ effectiveness in modern primary care populations.

## Data Availability

Dr. Liss had full access to all the data in the study and takes responsibility for the integrity of the data and the accuracy of the data analysis.

## Acknowledgments

We thank Jonna Peterson, MLIS (Northwestern University Feinberg School of Medicine, Galter Health Sciences Library and Learning Center) for her support and assistance throughout the study search and selection process, including the design and conduct of the MEDLINE search, support during the abstract review phase, and obtaining full text articles. Ms. Peterson received no compensation for her project contributions.

