## Supplement for "General Health Checks in Adult Primary Care: A Review"

**Ovid MEDLINE Search: Conducted January 19, 2021**

Part 1 of 3: Observational studies published between January 1, 2000 and December 31, 2020

1. physical examination/ and ((annual or gp or periodic or yearly or routine).ti. or ((primary adj2 (care or healthcare)) or primary health* or general practitioner? or general practice or family doctor? or family practice? or family physician?).ti,ab.)

2. (health screen* or health check* or healthcheck* or annual physical? or annual medical or medical check* or primary care check* or wellness check* or well care or wellcare or well woman or well visit?).ti.

3. ((annual or periodic or regular or routine or yearly) and (check* or health* exam* or health evaluation? or medical exam* or physical? exam* or wellness check* or gp visit? or physician? visit? or doctor? visit? or office visit?)).ti.

4. ((annual or yearly) adj2 (medical? or physical?)).ti.

5. ((annual or yearly) and visit?).ti.

6. (preventive? and (care check* or checkup? or check-up? or visit? or exam* or family doctor? or gp or family physician? or general practitioner?)).ti.

7. (general health screening or periodic health evaluation*).ti,ab.

8. or/1-7.

9. exp primary health care/

10. family practice/

11. physicians, primary care/

12. general practice/

13. physicians, family/

14. general practitioners/

15. exp outpatient clinics, hospital/

16. ambulatory care/

17. exp ambulatory care facilities/

18. exp community health services/

19. exp community health centers/

20. or/9-19

21. 8 and 20

22. (quasi-experiment* or natural experiment*).ti,ab.

23. (comparative study or evaluation studies or multicenter study).pt.

24. (review or systematic review).pt.

25. observational study.pt.

26. Epidemiologic studies/

27. exp case control studies/

28. exp cohort studies/

29. Case control.tw.

30. (cohort adj (study or studies)).tw.

31. Cohort analy$.tw.

32. (Follow up adj (study or studies)).tw.

33. (observational adj (study or studies)).tw.

34. Longitudinal.tw.

35. Retrospective.tw.

36. Cross sectional.tw.

37. Cross-sectional studies/

38. evaluation studies/ or evaluation studies as topic/ or program evaluation/ or validation studies as topic/ or (preadj5 post- or (pretest adj5 posttest) or (program* adj6 evaluat*)).ti,ab. or (effectiveness or intervention).ti,ab.

39. Comparative studies/

40. Follow-up studies/

41. prospective$.tw.

42. or/22-41.

43. exp animals/ not humans/

44. 42 not 43.

45. 21 and 44

46. (2000* or 2001* or 2002* or 2003* or 2004* or 2005* or 2006* or 2007* or 2008* or 2009* or 2010* or 2011* or 2012* or 2013* or 2014* or 2015* or 2016* or 2017* or 2018* or 2019* or 2020*).dc,dp,ed,ep,yr.

47. 45 and 46

48. Mass Screening/

49. Systematic risk assessment*.tw.

50. Case finding.tw.

51. ((screen* or assess* or test* or diagnos* or surveill* or identifi* or prevelence or incidence*) adj10 (structured or systematic or organised or organized or opportunistic or random)).tw.

52. Risk Assessment/

53. (risk* adj3 assess*).tw.

54. or/48-53

55. Primary Prevention/

56. 54 and 55

57. 8 or 56.

58. 57 and 20

59. 58 and 44

60. 59 and 46

Ovid MEDLINE Search, Part 2 of 3: Randomized trials of general health checks published between January 1, 2018 and December 31, 2020*

1. physical examination/ and ((annual or gp or periodic or yearly or routine).ti. or ((primary adj2 (care or healthcare)) or primary health* or general practitioner? or general practice or family doctor? or family practice? or family physician?).ti,ab.)

2. (health check* or healthcheck* or annual physical? or annual medical or medical check* or primary care check* or wellness check* or well care or wellcare or well woman or well visit?).ti.

3. ((annual or periodic or regular or routine or yearly) and (check* or health* exam* or health evaluation? or medical exam* or physical? exam* or wellness check* or gp visit? or physician? visit? or doctor? visit? or office visit?)).ti.

4. ((annual or yearly) adj2 (medical? or physical?)).ti.

5. ((annual or yearly) and visit?).ti.

6. (preventive? and (care check* or checkup? or check-up? or visit? or exam* or family doctor? or gp or family physician? or general practitioner?)).ti.

7. ((multifactor* or multi-factor*) adj5 prevent*).ti,ab.

8. (multiphasic adj2 (screening or test* or check*)).ti,ab.

9. comprehensive health test.ti,ab.

10. general health screening.ti,ab.

11. multiphasic screening/

12. ((diet or smoking or exercise or lifestyle or weight reduction or physical activity) and (screen* or check?) and (prevention or preventive or preventative)).ti,ab,hw.

13. or/1-12

14. mass screening/

15. ((general or prevent* or systematic or annual or yearly or periodic or regular or routine) adj5 (screen* or check? or checkup? or check-up?)).ti,ab.

16. (health check* or health screen*).ti,ab.

17. or/14-16

18. exp primary health care/

19. family practice/

20. physicians, primary care/

21. general practice/

22. physicians, family/

23. general practitioners/

24. exp outpatient clinics, hospital/

25. ambulatory care/

26. exp ambulatory care facilities/

27. exp community health services/

28. exp community health centers/

29. ((primary or communit*) adj5 (care or health*)).ti,ab.

30. (family practi* or family doctor* or family physician* or gp* or general practi*).ti,ab.

31. ((outpatient? or ambulatory) adj2 (care or healthcare or clinic? or service? or facilit*)).ti,ab.

32. or/18-31

33. 17 and 32

34. 13 or 33

35. exp randomized controlled trial/

36. controlled clinical trial.pt.

37. randomi#ed.ti,ab.

38. placebo.ab.

39. randomly.ti,ab.

40. Clinical Trials as topic.sh.

41. trial.ti.

42. or/35-41

43. exp animals/ not humans/

44. 42 not 43

45. 34 and 44

46. (2018* or 2019* or 2020*).dc,dp,ed,ep,yr.

47. 45 and 46.

* This date range represents the period since this MEDLINE search was conducted for a prior Cochrane review of general health checks (Krogsboll LT, Jorgensen KJ, Gotzsche PC. General health checks in adults for reducing morbidity and mortality from disease. *Cochrane Database Syst Rev*. 2019;1:CD009009.)

Ovid MEDLINE Search, Part 3 of 3: Randomized trials of systematic risk assessment for primary prevention of cardiovascular disease published between January 1, 2015 and December 31, 2020**

1. exp Cardiovascular Diseases/

2. cardio*.tw.

3. cardia*.tw.

4. heart*.tw.

5. coronary*.tw.

6. angina*.tw.

7. ventric*.tw.

8. myocard*.tw.

9. pericard*.tw.

10. isch?em*.tw.

11. emboli*.tw.

12. arrhythmi*.tw.

13. thrombo*.tw.

14. atrial fibrillat*.tw.

15. tachycardi*.tw.

16. endocardi*.tw.

17. (sick adj sinus).tw.

18. exp Stroke/

19. (stroke or strokes).tw.

20. cerebrovasc*.tw.

21. cerebral vascular.tw.

22. apoplexy.tw.

23. (brain adj2 accident*).tw.

24. ((brain* or cerebral or lacunar) adj2 infarct*).tw.

25. exp Hypertension/

26. hypertensi*.tw.

27. peripheral arter* disease*.tw.

28. ((high or increased or elevated) adj2 blood pressure).tw.

29. exp Hyperlipidemias/

30. hyperlipid*.tw.

31. hyperlip?emia*.tw.

32. hypercholesterol*.tw.

33. hypercholester?emia*.tw.

34. hyperlipoprotein?emia*.tw.

35. hypertriglycerid?emia*.tw.

36. exp Arteriosclerosis/

37. exp Cholesterol/

38. cholesterol.tw.

39. coronary risk factor*.tw.

40. Blood Pressure/

41. blood pressure.tw.

42. or/1-41

43. Mass Screening/

44. Systematic risk assessment*.tw.

45. Case finding.tw.

46. ((screen* or assess* or test* or diagnos* or surveill* or identifi* or prevelence or incidence*) adj10 (structured or systematic or organised or organized or opportunistic or random)).tw.

47. Risk Assessment/

48. (risk* adj3 assess*).tw.

49. or/43-48

50. Primary Prevention/

51. 42 and 49 and 50

52. randomized controlled trial.pt.

53. controlled clinical trial.pt.

54. randomized.ab.

55. placebo.ab.

56. drug therapy.fs.

57. randomly.ab.

58. trial.ab.

59. groups.ab.

60. 52 or 53 or 54 or 55 or 56 or 57 or 58 or 59

61. exp animals/ not humans.sh.

62. 60 not 61

63. 51 and 62

64. (2015* or 2016* or 2017* or 2018* or 2019* or 2020*).dc,dp,ed,ep,yr.

65. 63 and 64

** This date range represents the period since this MEDLINE search was conducted for a prior Cochrane review of systematic risk assessment for primary prevention of cardiovascular disease (Dyakova M, Shantikumar S, Colquitt JL, et al. Systematic versus opportunistic risk assessment for the primary prevention of cardiovascular disease. *Cochrane Database Syst Rev*. 2016(1):CD010411.)
